# Investigation of risk factors for congenital hypothyroidism (CH) using a population-based case-control study as part of the 2015-2018 screening program in Kohgiluyeh and Boyer Ahmad province in southwestern Iran

**DOI:** 10.1101/2024.04.27.24306474

**Authors:** Hedayatullah Jamali, Saied Bokaie, Ali Reza Bahonar, Hessameddin Akbarein, Reza Ramezani

## Abstract

Congenital hypothyroidism (CH) is a lack of thyroid hormone at birth, crucial for brain development. A nationwide screening program has been implemented in Iran since 2005, resulting in over 12 million infants being screened and more than 30,000 patients diagnosed and treated. The incidence of CH in Iran is 2.7 per thousand live births, which is higher compared to the global incidence of 1 in 3,000 to 4,000. The incidence of CH in Kohgiluyeh and Boyer Ahmad province is significantly higher than the national and global incidence. Prognostic factors including twins, the season of birth, puberty, jaundice at birth, birth weight, gestational age, anemia and goiter of the mother, type of delivery, father’s education, and smoking status are significantly associated with CH. Genetic, climatic, and environmental factors also play a role in the development of CH. Congenital hypothyroidism (CH) is one of the most common causes of intellectual disability, which can be prevented if diagnosed and treated on time. We aimed to investigate some related risk factors for CH in infants born in Kohgiluyeh and Boyer Ahmad province.

**Results:** This was a population-based case-control study conducted on 270 infants. The study population included infants who were born between 2015 and 2018 and were subjected to a screening program for CH. In general, 135 infants diagnosed with CH who were confirmed by a specialist and had a medical record were considered as the case group and the rest (135 infants) who were healthy were considered as controls. Patients and control infants were matched in a one-to-one ratio. Information was extracted from the Sib Health Integrated System (http://sib.yums.ac.ir). Regression analysis using the logistic regression method was performed on data collected from a sample of 270 infants and SPSS software version 24 was used to analyze the data with P<0.05 considered significant. Ethical considerations were addressed by obtaining approval from the ethics committee of Yasuj University of Medical Sciences under ethical number IR.YUMS.REC.1397.136 and holding preliminary meetings with authorities, health, and medical personnel to discuss different tasks, collaboration, and completion of checklists.

**Conclusion:** The results of the study show that among the cases studied, 3 factors, weight and height of the infant at birth and a family history of the disease in the infant could be the main risk factors for hypothyroidism (CH) in this province. Other factors such as birth order (rank), maternal age, maternal weight and height, age and mode of delivery, history of medication and iodized salt intake in the mother, diseases of the mother and father, and familial relationship of the parents were not observed to have a statistically significant association with CH. further studies are needed to analyze the results of the present study to establish the causality of these associations with greater certainty.

## Introduction

Congenital hypothyroidism is defined as a lack of thyroid hormone at birth, which is crucial for brain development. (1).

Congenital hypothyroidism (CH)^1^ is one of the most common causes of intellectual disability, which can be prevented if diagnosed and treated in time (2).

Congenital hypothyroidism is a disease that can cause severe mental retardation and developmental delay. However, timely diagnosis and treatment of infants with this disease can effectively prevent complications and delays in related development (3-4).

Congenital hypothyroidism is one of the most common preventable causes of mental retardation in children. Congenital hypothyroidism screening is one of the most cost-effective tools for preventing mental retardation in the population. CH has been part of newborn screening since the 1970s. (5-6).

Congenital hypothyroidism (CH) is a disorder that is very common in premature infants and is caused by maternal factors, complications before and around birth, genetic abnormalities, thyroid abnormalities, as well as side effects of drugs and therapeutic procedures. Because of this, prevention is not completely achievable. CH appears clinically in several distinct forms: primary, permanent or transient, and secondary. Their causes and consequences have little similarity. Therefore, accurate diagnosis and differentiation between subtypes of CH is very important to plan an effective treatment. (7).

Transient congenital hypothyroidism (CH) refers to a temporary deficiency of thyroid hormone, characterized after birth by low thyroxine (T4) and high thyrotropin (TSH), which later recovers and thyroxine production recovers, usually in The first few months of infancy. Approximately 17–40% of children diagnosed with CH through newborn screening programs (NBS)^2^ are later found to have transient hypothyroidism. Causes of transient CH include prematurity, iodine deficiency, maternal thyrotropin receptor-blocking antibodies, maternal use of antithyroid drugs, maternal or neonatal iodine exposure, loss of function mutations, and hepatic hemangiomas. Classic clinical signs and symptoms of CH are usually absent immediately after birth in the vast majority of infants due to temporary maternal thyroxine protection. NBS has been largely successful in preventing mental retardation by early detection of CH by performing thyroid function tests in infants with abnormal screening results. (8).

Congenital hypothyroidism (CH) is the most common endocrine disorder in children. Deprivation of thyroid hormone not only causes mental retardation, but also growth retardation (9-10).

Congenital hypothyroidism (CH) occurs in approximately 1:2000 to 1:4000 infants. Clinical manifestations are often subtle or absent at birth. This is probably due to some of the mother’s thyroid hormones crossing the placenta, while many infants make their thyroid. Common symptoms include decreased activity and increased sleep, difficulty feeding, constipation, and prolonged jaundice. On examination, common signs include myxedematous complexion, large fontanelles, macroglossia, distended abdomen with umbilical hernia, and hypotonia. CH is classified into permanent and transient forms, which in turn can be divided into primary, secondary, or environmental causes (11).

And if it is not diagnosed and treated in time, it can have devastating consequences in neurodevelopment. While newborn screening has virtually eradicated intellectual disability from severe congenital hypothyroidism in the developed world, more rigorous screening strategies have led to increased detection of mild congenital hypothyroidism. Recent studies provide conflicting evidence regarding the potential neurodevelopmental risks of mild congenital hypothyroidism, highlighting the need for further research to determine the risks these patients face and whether they are likely to benefit from treatment. he does. Furthermore, while the apparent incidence of congenital hypothyroidism has increased in recent decades, the underlying cause remains elusive in most cases (12).

The national screening program for neonatal hypothyroidism in Iran has been integrated into the country’s health system since 1384, and so far more than 12 million infants have been screened and more than 30,000 patients (suffering from both temporary and permanent types) have been diagnosed. and have been treated. Currently, the incidence of the disease is 2.7 per thousand live births (temporary and permanent), which is a sign of the high incidence of the disease in the country. The incidence of congenital thyroid hypothyroidism in Kohgiluyeh and Boyer Ahmad province is significantly higher than the national and global incidence according to the screening statistics conducted in the province. The national incidence is 1 in 1,000 and the global incidence is 1 in 3,000 to 4,000. (13).

According to the statistics available in the health centers of Kohgiluyeh and Boyer Ahmad province, this index is equal to 6.5 per thousand live births. The prevalence of hypothyroidism in infants during the years 2015 to 2018 in Kohgiluyeh and Boyer Ahmad province is about 5 per thousand live births.

Identifying the risk factors related to hypothyroidism in newborns, especially the preventable risk factors, and the epidemiological, clinical, and paraclinical investigation of the results of the national screening plan for hypothyroidism in newborns can be an effective measure in promoting the implementation of the program and reducing the incidence rate by carrying out effective interventions. And the spread of this disease at the level of this province and country.

According to the official statistics of the Ministry of Health, in recent years, the rate of incidence and prevalence of this disease in the country and the province of Kohgiluyeh and Bairahmad has been increasing, so that this province in the southwest of the country has the highest rate of the disease per thousand live births. Therefore, considering the importance of the issue and the lack of comprehensive study and research in Kohgiluyeh and Boyer Ahmad province, and considering the increase in the incidence rate and complications caused by this disease in this province and the country in recent years; This study was planned and carried out in this province, which examines the risk factors of hypothyroidism, as well as the epidemiological and clinical status of patients with hypothyroidism over several years in Kohgiluyeh and Bairahmad province. In a two-part checklist, demographic information and information related to the characteristics related to the disease were collected and analyzed. The location of Kohgiluyeh and Boyer Ahmad province is shown in Figure 1.

**Figure 1.**
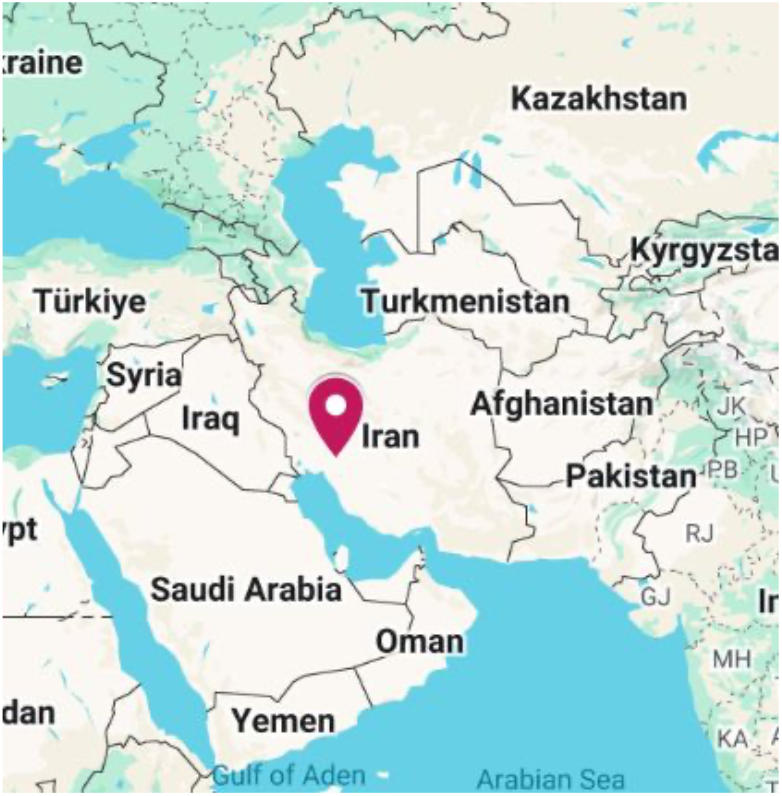
The location of Kohgiluyeh and Boyer Ahmad province.

Clinical complications of congenital hypothyroidism, such as brain disorders, are very subtle and cannot be detected during infancy. They are detectable when it is too late for treatment or prevention. General newborn screening is effective in diagnosing congenital hypothyroidism and starting early treatment. Early diagnosis of patients, while creating beneficial effects for patients and increasing the quality of life, reduces the long-term costs of the health system (14).

The most important prognostic factors that had a significant effect on congenital hypothyroidism include twins, the season of birth, puberty, jaundice at birth, birth weight, gestational age, anemia and goiter of the mother, gestational age, type of delivery, father’s education and status smoking and consanguinity. In most cases, the associations reported by logistic regression were stronger than propensity score analysis, although the differences were not statistically significant. However, future prospective studies are needed to test these findings and hypotheses and investigate the true impact of these potential risk factors on congenital hypothyroidism (15-17).

One of the most preventable causes of mental and developmental retardation is congenital hypothyroidism (CH) (5,18).

Despite significant progress in the management and treatment of congenital hypothyroidism, this disorder continues to cause major neurodevelopmental deficits in infants. Congenital hypothyroidism (CH) originates from several factors, so eliminating them can help reduce the prevalence of the disease (19).

Perinatal factors (gestational thyroid dysfunction, gestational diabetes, etc.) are associated with congenital hypothyroidism (CH). (20).

Congenital hypothyroidism (CH) is one of the main causes of mental retardation in infants. Also, this disease is related to genetic, climatic, and environmental factors. (6,21-22).

Studies show that the occurrence rate of CH in Iran is very high. (13,23).

The apparent incidence of congenital hypothyroidism has more than doubled in recent years due to several factors, including more comprehensive diagnostic criteria, changing demographics, and increased survival of preterm infants. The greatest increase occurred in patients with mild disease, many of whom have an atopic thyroid gland. Congenital hypothyroidism may be transient or persistent, but the natural history cannot be accurately predicted at diagnosis. In preterm infants, who are particularly vulnerable to hypothyroidism, the increase in thyroid-stimulating hormone may be delayed and therefore detected only by routine follow-up screening. Although newborn screening has virtually eradicated mental retardation due to congenital hypothyroidism in parts of the world, new information continues to be collected and new questions are raised about the diagnosis, physiology, and optimal management of this disorder (24).

Considering that differences in laboratory tests may cause changes in detection frequencies, there is a need for more careful investigation in this process to avoid the consequences of false positive and false negative results (25).

The economic burden of mental retardation caused by CH remains an important public health challenge in countries without routine newborn screening (NBS) programs. The health burden of CH remains high even in countries with well-developed NBS (26).

The performance of the newborn screening program for congenital hypothyroidism in Iran is favorable with a coverage of over 95%. Considering the high rate of recall and the incidence of CH, it is necessary to monitor the screening program in the country and also conduct more studies to determine the main risk factors for the high rate of recall and incidence of this congenital error. (27).

The main cause of CH worldwide, especially in Africa and Asia, remains iodine deficiency, which leads to goiter, deleterious forms of mental retardation, and in fetal life - stillbirth or spontaneous abortion or congenital malformations (cretinism). (28).

Congenital hypothyroidism occurs in approximately 1 in 2,000 infants and can have devastating consequences on neurodevelopment if not diagnosed and treated early. While newborn screening has virtually eradicated intellectual disability from severe congenital hypothyroidism in the developed world, more rigorous screening strategies have led to increased detection of mild congenital hypothyroidism. Recent studies provide conflicting evidence regarding the potential neurodevelopmental risks of mild congenital hypothyroidism, highlighting the need for further research to determine the risks these patients face and whether they are likely to benefit from treatment. he does. Furthermore, while the apparent incidence of congenital hypothyroidism has increased in recent decades, the underlying cause remains elusive in most cases (12).

Recent studies have shown an increase in the incidence of congenital hypothyroidism in the last 2 or 3 decades in the world. The cause of this change is unknown, various studies in different provinces of the country show that the prevalence of CH is higher compared to the global level. Therefore, comprehensive and supplementary studies to identify related risk factors should be prioritized in health system research. (24,29,30,31,32,33,3,19)

Due to the importance of the subject and the lack of research in Kohgiluyeh and Boboyrahmad province, this study was planned and implemented in this province from 2014 to 2019 to investigate the demographic and pathological variables (clinical and risk factors) that have been less discussed in other studies; which I hope will open the way for some medical and sanitary problems in the society and this province.

## Method

In a retrospective case-control study, the researchers examined all infants diagnosed with hypothyroidism between 2015 and 2018 in the province of Kohgiluyeh and Boyar-Ahmad. The information for these infants was obtained from the health centers and the newborn screening laboratory in the region; the samples were selected and examined by census. To perform this study, for each sick infant, a healthy infant was selected that was similar in terms of sex, year of birth (age), and place of residence to avoid possible bias. They were selected using the appropriate checklist and referring to the family file, their information was filled in and they were analyzed for the risk factors of congenital hypothyroidism. Descriptive analysis methods such as mean and standard deviation for quantitative variables and percentage frequency for qualitative variables were used to describe the subjects’ information. The chi-square test was used to analyze the qualitative variables. In general, chi-square tests, T-tests, and single and multivariate regressions with calculated CI and OR were used to calculate and examine the relationship between the variable studies. The effect of independent variables Disease was examined using single and multivariate logistic regression models with control for possible confounding effects. The disease was investigated using single and multivariate logistic regression models with the control of possible confounding effects. The data with P < 0.5 were entered into multivariate analysis. Regression analysis in multivariate analysis using the logistic regression method was done with a backward elimination procedure for variables. Data analysis was done using SPSS software version 24 and P<0.05 was considered significant.

### Section snippets

#### Ethical issues

This study was approved by the ethics committee of Yasuj University of Medical Sciences under ethical number IR.YUMS.REC.1397.136. Before starting the study, several preliminary meetings were held with authorities, health and medical personnel. Issues including different tasks, collaboration, how to complete checklists were discussed. The written agreements of the related cases were referred by the university to the health networks of the subordinate cities of Kohgiluyeh and Boyer Ahmad province.

#### The findings

Out of 270 infants, the case and control groups were 135 (68 girls and 67 boys) and 135 (68 girls and 67 boys), respectively. In general, in this study, boys were slightly more than boys at 49.6% and girls at 50.4%. 162 of the people under study lived in the city (60%) and 108 people lived in the village (40%). The highest birth order (rank) among the people under study (controls and cases) was the first rank or one of the first infants in the family with 40%.And the lowest was observed with rank more than 4th rank or 4th infant and later with 3%. The most people among witnesses and cases were in May with 13%, November with 12%.,and September with 10%, respectively. The type of delivery in the people under study was natural in 60% of cases and the rest were delivered by cesarean section. Family history of hypothyroidism was observed in 11.5% of cases with 31 people in the patient group and 6.7% of cases with 18 people in the control group. In this opinion, there was not a statistically significant difference between the two groups of case and control. (P = 0.58). Among the patients under study, most of the cases were transient with 24.1% And the rest include permanent cases with 25.9% and those under treatment whose disease status has not yet been determined by a specialist doctor in terms of transient or permanent nature of the disease and have been treated at 6.3%., there was a statistically significant difference between the two groups of case and control. Among the group of patients alone, transient type with 48.2%, permanent type with 39.2%, and the rest who have been treated and the type of their disease has not yet been determined equal to 12.6% have been observed (P < 0.001).

In univariate analysis, weight of birth time with odds ratio [[OR=0.99; 95% confidence interval [CI: 0.99-1, P=0.008), height at birth with odds ratio [[OR=0.87; 95% CI [CI: 0.79-0.96, P=0.005), family history of hypothyroidism with odds ratio [[OR=1.938; 95% confidence interval [CI: 1.02-3.66, P=0.04); have shown statistically significant. No statistically significant association was found for other variables such as birth order (rank), maternal age, maternal weight and height, age and mode of delivery, history of medication and iodized salt use in the mother, maternal and paternal illnesses, and parents’ familial relationship with CH. In the multivariate analysis, none of the factors included in the model showed a significant relationship with neonatal hypothyroidism except for the infants’ height (odds ratio [[OR=0.875; 95% confidence interval [CI: 0.79-0.96, P=0.005)). The results of the above statistical tests are shown in Tables 1 to 4.

**Table 1.** Results of the independent t-test and descriptive statistic comparing the two groups of case and control based on the quantitative indicators of the study.

| Variable | t-test for Equality of Means |  | 95% Confidence Interval of the Difference |  | Sig. (2-tailed) | N | Range |  | Mean±SD |  |
| --- | --- | --- | --- | --- | --- | --- | --- | --- | --- | --- |
|  | t | df | Lower | Upper |  |  | Minimum | Maximum | Total | Control Case |
| Time of birth(age) (day) | 0.365 | 268 | -.39017 | 0.56795 | 0.715 | 270 | 3.00 | 19.00 | 4.41±1.99 | 4.45±1.91<br>4.37±2.07 |
| Weight of the infant at birth(gr) | 2.713 | 245.030<sup>1</sup> | 46.25339 | 291.46513 | 0.007 | 270 | 1050.00 | 4550.00 | 3161.96±517.41 | 3246.39±425.98<br>3077.53±584.47 |
| Height of the infant at birth(cm) | 3.019 | 237.652<sup>2</sup> | .39134 | 1.86051 | 0.003 | 270 | 33.00 | 60.00 | 49.39±3.10 | 49.95± 2.45<br>48.82±3.56 |
| Mother's age (Year) | 0.617 | 268 | -.97330 | 1.86219 | 0.538 | 270 | 17.00 | 49.00 | 30.37±5.90 | 30.60±6.01<br>30.15±5.81 |
| Mother's weight(kg ) | -0.559 | 268 | -3.34574 | 1.86649 | 0.577 | 270 | 45.00 | 109.00 | 70.69±10.86 | 70.32±11.01<br>71.06±10.72 |
| Mother's height (m) | 0.931 | 268 | -.85418 | 2.38751 | 0.353 | 270 | 110.00 | 174.00 | 158.29±6.76 | 158.68±6.16<br>157.91±7.31 |
| age of delivery (week) | 0.346 | 268 | -.27835 | .39687 | 0.730 | 270 | 31.00 | 44.00 | 38.82 ±0.40 | 38.85±1.17<br>38.80±1.60 |

<sup>1</sup> Levene's Test for Equality of Variances is significant so (Equal variances not assumed) were reported
<sup>2</sup> Levene's Test for Equality of Variances is significant so (Equal variances not assumed) were reported

Based on the information provided in Table 1-4 here are the key results:

Table 1 provides the results of the independent t-test and descriptive statistics comparing the quantitative variables between the case and control groups.

-Variables like Time of birth (age/day), Weight of the infant at birth (g), and Height of the infant at birth (cm) showed statistically significant differences between the case and control groups, as indicated by the p-values less than 0.05.

-For example, the mean Time of birth (age/day) was 4.45 days in the control group and 4.37 days in the case group, with no statistically significant difference (p=0.715).

Of course, by mentioning that we had matched both the case and control groups based on age and gender. Table 2:

**Table 2.** Results of the chi-square test and descriptive statistic comparing the two groups of case and control based on the qualitative indicators of the study.

| Variable |  | N |  | Percent |  | Chi-Square Tests | p-value for Chi-Square |
| --- | --- | --- | --- | --- | --- | --- | --- |
|  |  | Control | Case | control | Case |  |  |
| Gender | Male | 67 | 67 | 24.8% | 24.8% | Continuity Correction | 1.000 |
|  | Female | 68 | 68 | 25.2% | 25.2% |  |  |
| Place of birth | the city | 81 | 81 | 30.0% | 30.0% | Continuity Correction | 1.000 |
|  | the village | 54 | 54 | 20.0% | 20.0% |  |  |
| Birth Order <sup>1</sup>(rank) | Order 1 | 57 | 52 | 21.1% | 19.3% | Pearson Chi-Square | 0.901 |
|  | Order 2 | 50 | 49 | 18.5% | 18.1% |  |  |
|  | Order 3 | 19 | 25 | 7.0% | 9.3% |  |  |
|  | Order 4 | 5 | 5 | 1.9% | 1.9% |  |  |
|  | Order >4 | 4 | 4 | 1.5% | 1.5% |  |  |
| Mode of delivery | Natural | 90 | 90 | 33.3% | 33.3% | Continuity Correction | 1.000 |
|  | Cesarean section | 45 | 45 | 16.7% | 16.7% |  |  |
| History of medication in the mother | YES | 24 | 30 | 8.9% | 11.1% | Continuity Correction | 0.447 |
|  | NO | 111 | 105 | 41.1% | 38.9% |  |  |
| History of iodized salt use in the mother | YES | 96 | 103 | 35.6% | 38.1% | Continuity Correction | 0.407 |
|  | NO | 39 | 32 | 14.4% | 11.9% |  |  |
| Father's illnesses | anemia | 6 | 1 | 2.2% | 0.4% | Pearson Chi-Square | 0.131 |
|  | diabetes | 5 | 5 | 1.9% | 1.9% |  |  |
|  | NO | 124 | 129 | 45.9% | 47.8% |  |  |
| Mother's illnesses | anemia | 7 | 6 | 2.6% | 2.2% | Pearson Chi-Square | 0.675 |
|  | diabetes | 5 | 8 | 1.9% | 3.0% |  |  |

<sup>1</sup> The order in which a child is born to a mother, compared to the mother's other children, is known as the birth order. This number reflects the sequence of the child's arrival within the family.

|  |  |  |  |  |  |  |  |
| --- | --- | --- | --- | --- | --- | --- | --- |
|  | NO | 123 | 121 | 45.6% | 44.8% |  |  |
| Family history of the disease | YES | 18 | 31 | 6.7% | 11.5% | Continuity Correction | 0.058 |
|  | NO | 117 | 104 | 43.3% | 38.5% |  |  |
| Parents' familial relationship | 2nd degree | 13 | 29 | 4.8% | 5.9% | Pearson Chi-Square | 0.570 |
|  | 3rd grade | 19 | 42 | 7.0% | 8.5% |  |  |
|  | 4th degree | 18 | 41 | 6.7% | 8.5% |  |  |
|  | distant relative | 11 | 24 | 4.1% | 4.8% |  |  |
|  | non-relative | 74 | 134 | 27.4% | 22.2% |  |  |
| Season of disease | Spring | 32 | 33 | 11.9% | 12.2% | Pearson Chi-Square | 0.984 |
|  | Summer | 40 | 38 | 14.8% | 14.1% |  |  |
|  | Fall | 39 | 38 | 14.4% | 14.1% |  |  |
|  | winter | 24 | 26 | 8.9% | 9.6% |  |  |
| Type of disease | Transient | 0 | 65 | 0.0% | 24.1% | Pearson Chi-Square | 0.000(<0.001) |
|  | Permanent | 0 | 53 | 0.0% | 19.6% |  |  |
|  | Under treatment (unspecified type yet) | 0 | 17 | 0.0% | 6.3% |  |  |
|  | Healthy (no disease) | 135 | 0 | 50.0% | 0.0% |  |  |

The chi-square test was used to compare the distributions of various categorical variables between the case and control groups.

-For variables like Gender, Place of birth, Birth Order, Mode of delivery, History of medication in the mother, and History of iodized salt use in the mother, the p-values for the chi-square tests were all or 1.000 or > 0.05, indicating no statistically significant differences between the case and control groups.

-For Birth Order, the Pearson chi-square test was used, and the p-value was 0.901, again showing no significant difference.

In summary, the chi-square test results indicate no significant differences between the case and control groups for the categorical variables, while the t-test results show significant differences for some of the quantitative variables like birth time, weight, and height of the infant.

According to the results shown in Table 3, which are based on the univariate logistic regression test, an increase in height of one centimeter reduces the probability of hypothyroidism in babies to 0.875, indicating a protective effect. This effect is statistically significant as the confidence interval does not include 1. Even after including this variable in the multivariate logistic regression model, it remains statistically significant (p = 0.005).

**Table 3.** Relationship between congenital hypothyroidism and its risk factors using univariate logistic regression test.

| Variable | Exp(B)/(OR) | 95% C.I for EXP(B) |  | p-value |
| --- | --- | --- | --- | --- |
|  |  | Lower | Upper |  |
| Height of the infant at birth(cm) | 0.875 | 0.797 | 0.960 | 0.005 |
| Weight of the infant at birth(gr) | 0.999 | 0.999 | 1.000 | 0.008 |
| Birth Order (rank) |  |  |  | 0.902 |
| Birth rank (1) | 0.912 | 0.217 | 3.835 | 0.900 |
| Birth ran (2) | 0.980 | 0.232 | 4.140 | 0.978 |
| Birth ran (3) | 1.316 | 0.291 | 5.949 | 0.721 |
| Birth ran (4) | 1.000 | 0.156 | 6.420 | 1.000 |
| Mother's age (Year) | 0.987 | 0.948 | 1.028 | 0.536 |
| Mother's weight(kg) | 1.006 | 0.984 | 1.029 | 0.575 |
| Mother's height (m) | 0.983 | 0.948 | 1.019 | 0.353 |
| Age of delivery | 0.970 | 0.818 | 1.150 | 0.729 |
| Age of delivery (week) | 1.000 | 0.603 | 1.659 | 1.000 |
| History of medication in the mother | 1.321 | 0.726 | 2.406 | 0.362 |
| History of iodized salt use in the mother | 1.308 | 0.759 | 2.253 | 0.334 |
| Maternal illnesses |  |  |  | 0.679 |
| Maternal illnesses (1) | 0.871 | 0.285 | 0.871 | 0.285 |
| Maternal illnesses (2) | 1.626 | 0.517 | 1.626 | 0.517 |
| Paternal illnesses |  |  |  | 0.242 |
| Paternal illnesses (1) | 0.160 | 0.019 | 0.160 | 0.019 |
| Paternal illnesses (2) | 0.961 | 0.272 | 0.961 | 0.272 |
| Family history of the disease | 1.938 | 1.024 | 3.667 | 0.042 |
| Parents' familial relationship |  |  |  | 0.571 |
| Parents' familial relationship (1) | 1.518 | 0.677 | 3.403 | 0.311 |
| Parents' familial relationship (2) | 1.493 | 0.744 | 2.996 | 0.259 |
| Parents' familial relationship (3) | 1.576 | 0.779 | 3.188 | 0.206 |
| Parents' familial relationship (4) | 1.458 | 0.609 | 3.487 | 0.397 |

Similarly, a weight gain of one gram in infants leads to a 0.999 probability of hypothyroidism in infants, indicating a weak protective effect. This effect is also statistically significant, as the confidence interval excludes the value 1. After this variable was included in the multivariable logistic regression model, it remained statistically insignificant (p = 0.255).

On the contrary, a family history of hypothyroidism in infants increases the probability of this disease to 1.906, indicating that it is a risk factor. This effect is statistically significant because the confidence interval does not include the number 1. However, this variable is not statistically significant after being included in the multivariable logistic regression model, although the p-value is close to 0.05 (p = 0.056).

Table 3 also shows the relationship between congenital hypothyroidism and its risk factors using a univariate logistic regression test. Some notable findings:

Paternal illnesses (1) had an Exp(B)/OR of 0.160 with a p-value of 0.019, indicating a potentially significant risk factor.

Paternal illnesses (2) had an Exp(B)/OR of 0.961 with a p-value of 0.272, suggesting it is not a significant risk factor.

Family history of the disease had an Exp(B)/OR of 1.938 with a p-value of 0.042, indicating it may be a significant risk factor.

Table 4 shows the relationship between congenital hypothyroidism and its risk factors using a multivariable logistic regression test. Key results include:

**Table 4.** Relationship between congenital hypothyroidism and its risk factors using multivariable logistic regression test.

| Variable | Exp(B)/(OR) | 95% C.I for EXP(B) |  | p-value |
| --- | --- | --- | --- | --- |
|  |  | Lower | Upper |  |
| Height of the infant at birth(cm) | 0.872 | 0.792 | 0.960 | 0.005 |
| Weight of the infant at birth(gr) | 1.000 | 0.999 | 1.000 | 0.255 |
| Mother's height (m) | 0.995 | 0.956 | 1.036 | 0.807 |
| History of medication in the mother | 1.121 | 0.515 | 2.437 | 0.774 |
| History of iodized salt use in the mother | 1.219 | 0.683 | 2.177 | 0.503 |
| Paternal illnesses |  |  |  | 0.160 |
| Paternal illnesses(1) | 0.124 | 0.014 | 1.087 | 0.059 |
| paternal illnesses(2) | 0.762 | 0.200 | 2.907 | 0.690 |
| Family history of the disease | 1.906 | 0.985 | 3.689 | 0.056 |

The height of the infant at birth had an Exp(B)/OR of 0.872 with a 95% CI of 0.792-0.960 and a p-value of 0.005, suggesting it is a significant risk factor.

The weight of the infant at birth had an Exp(B)/OR of 1.000 with a 95% CI of 0.999-1.000 and a p-value of 0.255, indicating it is not a significant risk factor.

-Mother’s height had an Exp(B)/OR of 0.995 with a 95% CI of 0.956-1.036 and a p-value of 0.807, suggesting it is not a significant risk factor.

The history of medication in the mother had an Exp(B)/OR of 1.121 with a 95% CI of 0.515-2.437 and a p-value of 0.774, indicating it is not a significant risk factor.

## Discussion

There is little information about the factors affecting hypothyroidism in infants in Kohgiluyeh and Boyer Ahmad province. Therefore, this study was carried out to investigate the factors affecting the hypothyroidism of infants in Kohgiluyeh-Boboyrahmad province, located in the southwest of Iran. Congenital hypothyroidism is the most common congenital endocrine disorder worldwide. Approximately 80-85% of cases are caused by defects in thyroid development (dysgenesis), and the remaining 15-20% are due to errors in thyroid hormone biosynthesis (dysmorphogenesis). Its incidence rate is 1 out of 3000/4000 live births worldwide, but this incidence rate is in developing countries (34).

The annual incidence of hypothyroidism is 1:16,000 live births in the Netherlands, 1:2,000 to 1:3,000 live births in the United Kingdom, and 1:1,266 live births in Sri Lanka in the country’s surveyed population. Most of them show no clinical manifestations or symptoms in the first weeks of life. In short, infants will develop mental retardation if they are not diagnosed (56-58).

Based on Ford G et al’s 2014 review of current screening programs, approximately 71 percent of infants worldwide are not born in an area with an established NBS program, even though screening has been in place in developed countries for over five decades. Thus, the majority of infants with CHD worldwide are not detected and treated early, so the economic burden of retardation due to CHD remains a significant public health challenge(59).

The results of this research showed that out of 135 patients with congenital hypothyroidism, 67 people(49.6%) were boys and 68 people (50.4%) were girls. Considering that the gender ratio of those born in the province is about 103 to 100, it was expected that the gender ratio of the infected people would be in the same range;While,like the research done in Mexico; The percentage of female patients in that country has slightly exceeded that of boys (1). Although the gender ratio has changed in 9 provinces of the country, including Kohgiluyeh and Boyer Ahmad, in recent years, female gender has been introduced as a risk factor in most of the studies conducted in the field of studying the risk factors of congenital hypothyroidism.Of course, it is not possible to analyze the effect of the variables mentioned in this study due to the maching of the variables gender, age of the infant, place of residence and season(month) of birth in this study. However, the frequency of patients shows that girls may be more at risk of neonatal hypothyroidism in the region and this province. Although the cause is unclear and needs more research and investigations. However, in a study in Central Province, there was a significant relationship between gender and hypothyroidism. (23). In another study conducted as a case-control study by Ahmed Mahmoud Al-Muktader in the Egyptian province of Al-Fiyum, as well as in a study conducted in China, a statistically significant association between CHD and female gender was found, which could be due to the lack of matching in these studies (36, 35). However, in a study conducted by Seyyed Mahmoud Rizvani and colleagues in Gilan and another study in Henan Province, China, no significant association was found between gender and CHD (37, 60).

The study by ZhouJinfu et al in Fujian Province, China, showed that women aged 35 years or older and those who suffered from thyroid disease and/or diabetes mellitus during pregnancy had an increased risk of their offspring developing CH. Infants of the female gender, premature births, afterbirths, low birth weight, other birth defects, and those born as part of multiple births also had an increased risk of CH(43.(

A 2018 study by Yousef Alimohamadi and colleagues in Khuzestan province showed that there was a significant association between female gender and twins with CH. However, as in the present study, the relationship between consanguinity and CHD was not statistically significant according to the results of the multivariate analysis in this study(61.(

In many studies in the world,the decrease in iodine intake is known as one of the most important risk factors for congenital hypothyroidism (38, 17, 32). The amount of iodized salt consumed by mothers and Pregnant women in this study at the level of Kohgiluyeh and Boyer Ahmad province was on average above 70% and no statistically significant difference was observed between the two groups of cases and controls. However, in a case-control study in Khuzestan province, there was a significant relationship between gender (female) and hypothyroidism. (16). In this study, similar to a study in Mazandaran province in northern Iran, the prevalence of transient CH was higher than that of permanent CH (39). In this study, it was not possible to take a urine sample to measure excreted iodine. It is possible that other factors such as the type of salt consumed or its concentration of iodine and the way food is cooked cause the mother or her infant to not get enough iodine. Therefore, it is necessary in this regard as well. More extensive research should be done. In this study, 23% of the infants with congenital hypothyroidism had a history of thyroid diseases in their family members, especially their mothers,which to the univariate analysis, there was a significant statistical difference between the two groups. There was a witness, but in the multivariate analysis, this difference was not significant, like the study that Mr. Afkhamzadeh and his colleagues conducted in Kurdistan province in 2008 and 2009 (40).

A nested case-control study of a cohort of 2.5 million births in California found that iodine status was not associated with CHD in the US population, which is reassuring given the widespread maternal iodine deficiency in the US. Among infants in the neonatal intensive care unit (NICU), cases of CHD had higher blood iodine concentrations compared with controls, suggesting that excessive iodine exposure in the NICU may cause CHD. Monitoring iodine exposure from surgical procedures, imaging and iodine-containing disinfectants and considering iodine-free alternatives may be helpful(62).

In this study, the birth weight of infants has a statistically significant relationship with congenital hypothyroidism like most studies in the country and abroad (41,37,29,18,3,41,42).

However, some studies have not found this relationship to be significant (40).

A study also found that preterm infants, especially those with very low birth weight (VLBW), are at increased risk for congenital hypothyroidism (43). The findings of this research also show the importance of screening and early diagnosis of congenital hypothyroidism in premature infants for the care of these infants.

A study conducted by Bret Nolan DO and colleagues in California (USA) in 2024 recommends that the incidence of congenital hypothyroidism in very preterm (VPT) or very low birth weight (VLBW) infants is unusual and requires thyroxine therapy(63).

Also, in this research, a statistically significant relationship was observed between the Height of birth of infants and congenital hypothyroidism. Similar to the studies that were conducted inside or outside the country (44,45). In a study in Iraq, it was proven that shortness Height with various changes in skeletal appearance is usually caused by juvenile hypothyroidism. Hypothyroidism in adolescents, especially girls, leads to various skeletal manifestations, including height reduction (46). However, studies in Iran and South Korea have not found this relationship to be significant (9,40).

In this study, family history has a statistically significant relationship with congenital hypothyroidism, especially the history of the mothers of newborns. Like most studies inside and outside the country (40, 47, 6, 49, 48).

In one study, it was found that maternal thyroid disease greatly increases the risk of neonatal/postnatal hypothyroidism (51,50).

However, some studies have not considered this relationship to be significant, especially the family history of the disease in mothers of newborns (52).

In another study conducted by Ms. Fariba Abbasi and colleagues in 2021 in northwestern Iran in East Azarbaijan province as part of a population-based case-control study of 340 infants sick and 340 healthy infants, family history, neonatal jaundice, gestational age at birth, and betadine use in pregnancy were introduced as risk factors for hypothyroidism in newborns(50.(

The data of this study in this province showed that about 48% of infants with primary CH had transient thyroid dysfunction. Our results clarify previous research by providing evidence on the incidence of CH. The rate of occurrence of CH as well as the transient type of CH in this province has been higher than those reported by other studies conducted in Iran and other regions of the world (13).

Other variables studied, including birth order (rank), maternal age, maternal weight and height, age and mode of delivery, history of medication and iodized salt use in the mother, maternal and paternal illnesses, and familial relationship of the parents had no significant association with congenital hypothyroidism in newborns, similar to studies within and outside the country (35,40,29,16,28,37). However, it has been seen in studies that had a statistically significant relationship with congenital hypothyroidism (10,50,20,51,53,21,35).

Studies conducted in China have shown that gestational blood pressure, gestational diabetes, thyroid disease during pregnancy, maternal age, premature birth, late delivery (age at birth), low birth weight infants, macrosomia, twins, and infants with malformations. Birth and infection were higher in the case group than in the control group and were statistically significantly associated with congenital hypothyroidism (3, 36, 29).

In a case-control study conducted by Yan, Xueqin et al. in 2023 in southern China with 255 case infants and 246 control infants, it was shown that thyroid dysfunction during pregnancy, gestational diabetes mellitus, and gestational age were the risk factors for neonatal hypothyroidism(20).

In another case-control study by Karcz, Karolina et al. from 2023 in Poland, conducted on 70 infants, it was found that newborns of mothers diagnosed with both gestational diabetes mellitus (GDM) and hypothyroidism had lower birth weight and fat mass than newborns of mothers without GDM or hypothyroidism. and mentioned that this result could be related to the high incidence of excessive weight gain during pregnancy in healthy mothers(64).

The results of the population-based case-control study in the province of Khorasan Razavi showed that time of birth(age), place of residence, maternal hypertension and depression, and consanguinity were the main risk factors for CH(65).

A study in East Azarbaijan province also revealed that the age of delivery had a statistically significant relationship with congenital hypothyroidism. In another study that was conducted in Hamadan, in addition to gestational age, anemia, and maternal goiter, Zeman type, The most important risk factors were maternal hypothyroidism and showed a significant relationship with CH (41,49).

Also, in a study that was conducted in Gilan, late delivery had a significant relationship with CH (37). A study of married couples in Khuzestan province also showed that the history of family marriage and urbanization had a statistically significant relationship with congenital hypothyroidism. is (54). A study in Ulaanbaatar, Mongolia showed that the main risk factors for primary congenital hypothyroidism are maternal age, birth weight, and gestational age (41).

Therefore, it is necessary to consider the above factors as risk factors in the control care of pregnant mothers, and if there is a history of hypothyroidism in the family or the infant is underweight or Stunting, it should be monitored by medical staff and the medical staff should be specially trained.

## Suggestions

As height and weight as well as family history are the most important risk factors for congenital hypothyroidism (CHT) in this study, expectant mothers are advised to pay particular attention to their care during pregnancy. Doctors, midwives, and healthcare workers should also be trained in this regard and pay special attention. It is also recommended to carefully assess the height and weight and family history, especially maternal illness, and record the results in their files to prevent the occurrence of CHT in time.

Coordination of diagnosis, management, and routine care not only optimizes patient outcomes but should also facilitate epidemiological studies of the disease. People with CH need to be monitored throughout their lives, especially in early childhood and during pregnancy (55).

it is suggested that more studies should be conducted at the country level to gain more certainty about the causality of these associations. Especially in the regions and provinces where the disease is more prevalent, it is necessary to conduct studies similar to the one mentioned above on the factors that influence this disease.

## limitation & Problems during project execution

Among the problems and limitations of this study is the incompleteness of the information and the incompleteness of Form No. 1 and 4 related to the screening of newborns and the care of patients with congenital hypothyroidism of newborns and the records of the patients; also, with the start of the pandemic of the Covid-19 disease and the involvement, and the participation of the health personnel of the province in this regard Collecting data to carry out the plan faced a lot of challenges, but despite the mentioned problems, by using the information received by contacting the families and parents of the patients or by using the search in the integrated health system (Sib; http://sib.yums.ac.ir); With the help and cooperation of health care workers and caregivers and the health team of comprehensive health service centers in the cities of in Kohgiluyeh and Boyer Ahmad province, the defect was removed and completed. Another limitation of this study is the lack of analysis of other potential risk factors such as genetic predisposition, environmental exposure, and iodine deficiency.

In this study, we were able to find suitable witnesses for each of the individuals in the case group so that the source of their selection was chosen according to the selection method of the case group and the individuals in the case group are representative of all cases in the population. In fact, this study is a population-based study with less possibility of selection bias. However, a small number of cases were excluded from the study due to migration and missing records based on the inclusion and exclusion criteria. However, we recognize that most case-control studies suffer from these problems, and no study is immune from the occurrence of systematic error or bias. Despite these limitations, this study used a number of potentially confounding variables such as gender, age, and place of birth by applying the matching method before analyzing the data, which increased the accuracy of the study and controlled for its effects.

## Conclusion

The results of the study show that among the cases studied, only 3 factors, weight and size of the infant at birth and a family history of the disease in the infant, maybe the main risk factors for CHD. Other factors such as age and mode of delivery, birth order (rank), mother’s age, mother’s weight and height, mother’s medication and iodized salt history, mother’s and father’s diseases, and Parents’ familial relationship were not statistically significant. However, further studies are needed to analyze the results of the present study and to ensure the causality of these associations.

## Data Availability

All data produced in the present study are available upon reasonable request to the authors.

## Acknowledgments

In the end, the authors consider it necessary to acknowledge the cooperation and assistance of the respected Health Vice-Chancellor of the University of Medical Sciences of Kohgiluyeh and Boyer Ahmad province, as well as the personnel of the health centers and health networks of the cities of Kohgiluyeh and Boyer Ahmad province, who helped us in carrying out the various stages of this research. helped and without their cooperation, this project would not have been possible, they expressed their sincere gratitude. This research is derived from a research project with the code of ethics IR.YUMS.REC.1397.136..

## Footnotes

1 Congenital hypothyroidism

2 Newborn Screening

